# Agentic Autodiscovery of Diastolic Dysfunction Phenotypes from Surface Electrocardiogram

**DOI:** 10.64898/2026.06.17.26355897

**Authors:** Ankush D. Jamthikar, Aditya Shanmugham, Sajeev Singh, Aditya Radhakrishnan, Jiawei Dong, Kameswari Maganti, Naveena Yanamala, Partho P. Sengupta

## Abstract

**Background:** Left ventricular diastolic dysfunction (LVDD) is a major determinant of heart failure (HF), yet its assessment relies on multiparametric echocardiography, limiting scalability. We previously demonstrated that generative artificial intelligence (AI) can synthesize tissue Doppler imaging (TDI) waveforms from the 12-lead ECG. The growing complexity of candidate architecture creates a need for automated model-discovery frameworks.

**Objectives:** To evaluate agentic AI-based auto-discovery for ECG-based LVDD assessment using either raw ECG or synthetic TDI waveforms.

**Methods:** Two attention-based agentic AI architectures were developed using an automated large language model-driven refinement framework that optimized transfer-learning and multimodal architectures through autonomous proposal, validation, and selection of candidate model configurations. Development was performed in 1,011 paired ECG-echocardiography studies and externally validated in 983 patients using two reference frameworks: (i) data-driven phenogroups and (ii) the 2025 ASE Diastolic Function Guidelines. External validation was performed in CODE-15% (n=219,567) for HF-related mortality and EchoNext (n=35,718) for structural heart disease associations.

**Results:** Despite the modest cohort size, the ECG-based agentic search achieved area under the receiver operating characteristic curve (AUCs) of 0.87 (95% CI: 0.85-0.89) and 0.83 (95% CI: 0.80-0.86) for phenogroup and guideline-based LVDD severity classification. Corresponding AUCs for the synthetic TDI-based model were 0.82 (95% CI: 0.80-0.85) and 0.80 (95% CI: 0.77-0.84), respectively. In large-scale external validation, both models stratified incident HF mortality with subdistribution hazard ratios ranging 5.5 to 9.5 (Gray’s p<0.001 for all). Time-dependent discrimination for incident HF mortality exceeded a publicly available convolutional neural network model (ECG2HF) (ΔAUC range: +0.14 to +0.20). Both models demonstrated consistent associations with structural heart disease outcomes.

**Conclusions:** Agentic auto-discovery enabled data-efficient assessment of LVDD from surface ECG by combining physiologically informed transfer learning with autonomous architecture optimization, achieving robust external generalizability. This approach may facilitate broader access to diastolic function assessment beyond conventional echocardiography.

## Introduction

Left ventricular diastolic dysfunction (LVDD) represents a latent disturbance in cardiac structure and function, characterized by abnormalities in myocardial relaxation, filling, or distensibility.^1^ Despite its central role in the development and progression of heart failure (HF),^2^ assessment of LVDD remains challenging. Approaches to LVDD phenotyping generally fall into two broad categories: conventional guideline-based frameworks and data-driven phenotyping strategies. Expert consensus guidelines, most recently the 2025 American Society of Echocardiography (ASE) diastolic function framework, provide a clinically grounded, heuristic-driven approach that integrates multiple echocardiographic parameters to classify diastolic function severity.^3^ In parallel, data-driven phenogroups, derived from unsupervised machine learning capture latent diastolic subtypes defined by patterns of co-occurring echocardiographic features and linked to distinct outcome trajectories.^4–6^ However, echocardiography is resource-intensive, creating a substantial scalability barrier, particularly in settings such as population screening, serial surveillance, or resource-limited environments.

The 12-lead electrocardiogram (ECG), by contrast, has near-universal availability and low cost.^7^ Myocardial diastolic function is physiologically encoded within surface ECG signals and AI models have demonstrated the capacity to detect LVDD from the ECG alone.^8,9^ More recently, our group has shown that generative AI models can synthesize patient-specific tissue Doppler imaging (TDI) waveforms directly from the 12-lead ECG, producing physiologically interpretable myocardial velocity profiles analogous to those obtained by Doppler echocardiography.^10^ At the same time, the rapid expansion of deep learning architectures—including choices related to feature encoders, attention mechanisms, multimodal fusion strategies, latent representations, training objectives, and optimization schemes—has made model development increasingly dependent on labor-intensive experimentation.^11^ Agentic AI offers a complementary paradigm in which candidate architectures are iteratively proposed, evaluated, and refined, enabling efficient exploration of large model-design spaces while reducing manual design bias. More broadly, large language models-based AI agents have been recently proposed as autonomous computational systems capable of coordinating complex research workflows, including hypothesis generation for candidate architecture selection, data analysis, model optimization, interpretation and discoveries.^12–14^ Together, these advances create an opportunity for knowledge synthesis using data-efficient and scalable ECG-based diastolic phenotyping.

Accordingly, this study sought to develop and compare two attention-based agentic AI architectures—one applied to raw 12-lead ECG waveforms and one to ECG-derived synthetic TDI waveforms—for diastolic dysfunction severity assessment using two reference standards: guideline-based categories (2025 ASE Criteria^3^) and data-driven diastolic phenogroups.^4,6^ Secondary objectives were to evaluate the prognostic equivalence of both models for incident heart failure and structural heart disease, and to assess whether model agreement and calibration were consistent across both classification frameworks in large external validation cohorts (**Central Illustration**).

## Method

### Study Population and Design

For agentic model training and validation, we included paired ECG and echocardiography data from 1,994 patients. This included a training cohort from 1,011 patients from a previously described multicenter study conducted at West Virginia University, the Icahn School of Medicine at Mount Sinai, and Windsor Cardiac Centre between 2017 and 2020 for developing an AI-ECG model of diastolic dysfunction^15^. The external validation cohort included 983 from the outpatient clinic who had an ECG and echocardiogram performed at Rutgers Robert Wood Johnson Medical School (New Brunswick, NJ, USA), collected between 2021 and 2023. Eligible adults (≥18 years), in whom a comprehensive echocardiogram was performed for LV systolic and diastolic function assessment, excluding those with paced rhythms, arrhythmias, non-interpretable ECG signals, a prosthetic valve in mitral position or mitral valve repair, congenital heart disease, cardiac transplantation, severe mitral annular calcification, any hemodynamic instability and inadequate or poor echocardiography quality.

Two external datasets, CODE-15%^16^ and EchoNext^17,18^, were used for additional model evaluation. The CODE-15% dataset^16^ is a stratified 15% sample of the Telehealth Network of Minas Gerais, Brazil, comprising primary-care and outpatient 12-lead ECG recordings collected between 2010 and 2016. Of 325,959 participants (median follow-up 3.4 years; interquartile range, QR: 2.1-5.1), 106,387 were excluded for incomplete follow-up or absent mortality data, yielding a final prognostic cohort of 219,567 patients. Cause of death was adjudicated from national death registries and classified as cardiac—subdivided into myocardial infarction-related, heart failure-related, and other cardiac causes—or non-cardiac. This dataset was used to assess the ability of both models to stratify heart failure-related mortality risk across two classification frameworks.

The EchoNext dataset comprises 100,000 ECGs from Columbia University Irving Medical Center and Allen Hospital, retrospectively collected from adults (age ≥18 years) who underwent paired 12-lead ECG and transthoracic echocardiography within a one-year interval between 2008 and 2022, with waveforms sampled at 250 Hz. After retaining first-recorded ECGs and excluding those with inadequate signal quality for synthetic TDI generation, 35,718 patients formed the final structural heart disease association cohort. This dataset was used to compare the association of ECG-based and TDI-based model-predicted risk classifications with a broad spectrum of echocardiographically confirmed structural heart disease outcomes—including valvular disease, ventricular dysfunction, elevated pulmonary pressures, and pericardial effusion.

Institutional review board approval was obtained from each participating site,^15^ and written informed consent was obtained from all prospective participants. The EchoNext and CODE-15% datasets are publicly available and were used in accordance with applicable data-use agreements.

### ECG and Echocardiographic Acquisition

In the training cohort, paired ECG and echocardiographic data were obtained on the same day, preferably within three hours of each other. In the validation cohort, ECGs were obtained on the patient’s arrival date, with paired echocardiograms recorded within 60 days (median 15 days; IQR 14-21 days). All recordings underwent band-pass filtering (0.5-40 Hz), quality control, and were segmented from R-peak to R-peak and normalized to a canonical 512-sample window. Two-dimensional transthoracic echocardiography was performed in all study participants according to standard clinical protocols, with measurements including diastolic function parameters, left ventricular systolic function, and cardiac chamber quantification performed in accordance with ASE recommendations.^19^ Diastolic dysfunction grading was performed in accordance with the 2025 ASE Diastolic Function Guidelines.^3,20^ For the EchoNext and CODE-15% external datasets, ECG waveforms were sampled at 250 Hz and 400 Hz respectively (**Supplemental Methods, Section S1**).^16,18^

### Generating Synthetic Tissue Motion

Synthetic TDI waveforms were generated from standard 12-lead ECGs using a previously validated generative adversarial network (GAN) model leveraging the electromechanical coupling between cardiac electrical activity and mechanical motion.^10^ Individual ECG cycles were standardized and processed by the GAN to reconstruct corresponding myocardial velocity waveforms. Synthetic TDI waveforms were generated across multiple cardiac cycles and averaged to produce a single representative waveform for each patient. External validation demonstrated strong correlation between synthetic and real TDI waveforms (repeated-measures r = 0.90, P < 0.0001), with clinical utility demonstrated for both diastolic and systolic dysfunction detection (AUC = 0.80 and 0.81, respectively).^10^

### Diagnostic Utility of Directly Acquired TDI Waveforms

Before evaluating ECG-derived synthetic TDI waveforms, we sought to determine whether the native TDI waveform alone contained sufficient information for diastolic classification. A convolutional neural network trained on directly acquired TDI waveforms achieved AUROCs of 0.81 and 0.82 for phenogroup and guideline-based LVDD classification, respectively (**Supplemental Figure 1; Supplemental Table 1**). These findings establish that the TDI waveform is a rich carrier of diastolic information and support the premise that faithful ECG-based synthesis of TDI signals can enable echocardiography-independent diastolic phenotyping.

### Model Development

To enable agentic auto-discovery of diastolic phenotypes from the surface ECG, we developed an automated multimodal AI framework in which a large language model (LLM)-driven refinement loop iteratively explored and optimized model architecture without manual intervention, guided by empirical performance signals.

#### (i) Input Modalities and Model Configurations

Each patient contributed up to three inputs and two demographic variables: a resting 12-lead surface ECG, an ECG-derived synthetic TDI velocity waveform generated using a previously validated generative AI model,^10^ and an ECG-derived myocardial displacement waveform reconstructed from the same generative framework. Age and biological sex were included as demographic covariates. Two model configurations were developed: (1) an ECG-based model using the 12-lead ECG with age and sex, and (2) a TDI-based model using both synthetic TDI-based waveforms with age and sex. Both model configurations were trained under each classification framework independently—once using phenogroup labels and once using guideline-based labels­^3,20^—to enable direct comparison across frameworks.

#### (ii) Attention-Based Architecture

Both configurations shared a unified transformer-based multimodal fusion architecture. Each modality was processed through a dedicated encoder compressing raw signal data into a compact learned representation, and the resulting representations were integrated through cross-modal attention layers to produce a final diastolic phenotype classification. The architecture was designed to remain functional when one or more inputs are unavailable in clinical use, as ECG and echocardiographic data are rarely co-acquired in routine care. This was achieved through a combinatorial modality-masking procedure, applicable across all model configurations, in which the model was simultaneously trained on all possible subsets of its inputs—ensuring well-calibrated predictions regardless of which modalities are available for a given patient. (**Supplemental Methods, Section S2**)

#### (iii) Agentic Auto-Discovery: LLM-Driven Model Refinement

The central feature of this framework is an automated, closed-loop agentic discovery process in which an LLM autonomously directed model development without human intervention at each iteration. Rather than relying on manual tuning, the agent was conditioned on the full history of prior training iterations, including observed performance metrics and diagnostic profiles, and tasked with proposing targeted modifications to model architecture and training procedures. Each proposal underwent a multi-stage validation protocol before execution, including checks for data leakage, duplication of prior failed proposals, and a signal-to-noise criteria ensuring predicted performance gains were meaningful rather than within the range of chance variation. Accepted proposals were implemented, trained end-to-end, and either retained or reverted based on validation performance.

### Echocardiographic Phenogrouping and Guideline-Based Classification

Two reference frameworks were used for LVDD classification. Two complementary reference frameworks were used for LVDD classification. First, we employed a previously developed phenogrouping approach based on topological data analysis and deep neural network modeling, which integrates multiple echocardiographic measures of left ventricular structure and function into a patient similarity network.^4,6^ This agnostic, data-driven method assigns patients to one of four phenogroups representing progressive stages of cardiac dysfunction with distinct prognostic trajectories. Patients belonging to higher risk phenogroups exhibit substantially greater rates of heart failure hospitalization and mortality independent of traditional risk factors.

Second, diastolic dysfunction severity was classified according to the 2025 ASE Diastolic Function Guidelines^3^ using a published JavaScript code to automate the assessment^20^. Individuals lacking sufficient parameters for DD25 grading were classified as indeterminate as previously described. Specifically, studies missing E, A, and both e′ measurements were considered indeterminate.^21^ Diastolic function was classified when at least one e′ measurement and the remaining primary parameters were available. Consistent with DD25 criteria, individuals with normal e′ velocities, normal E/A and E/e′ ratios, but unavailable peak tricuspid regurgitant velocity (TRV), were also categorized as indeterminate. The availability of echocardiographic measurements is summarized in **Supplemental Table 2**. For analysis, graded cases were dichotomized into Normal/Grade 1 and Grade 2/3 groups.

Both frameworks were evaluated because they represent complementary paradigms for characterizing diastolic dysfunction. Phenogrouping provides an unsupervised, data-driven assessment of patient similarity, whereas guideline-based grading reflects contemporary expert consensus and current clinical practice.

### Outcome Associations

For mortality outcome assessment, heart failure-related death was the primary endpoint, with myocardial infarction-related, other cardiac, and non-cardiac deaths treated as competing events in the CODE-15% dataset.^16^ For structural heart disease outcome assessment, both models were assessed against echocardiographically confirmed structural heart disease labels from the EchoNext dataset,^17,18^ including left ventricular hypertrophy (LVH; maximum wall thickness ≥1.3 cm), LV systolic dysfunction (EF <50%), RV systolic dysfunction, pulmonary hypertension (PASP ≥45 mmHg), valvular heart disease and (moderate or greater aortic stenosis, mitral or tricuspid regurgitation).

### Statistical Analysis

Continuous variables are presented as median (IQR) and categorical variables as frequencies (%). Group comparisons were performed using t-tests or one-way ANOVA for normally distributed variables, Mann-Whitney U or Kruskal-Wallis tests for non-normally distributed variables, and chi-square tests for categorical variables.

Model discrimination was quantified using area under the receiver operating characteristic curve (AUC) with 95% confidence intervals calculated using the DeLong method; each model was retrained 20 times under independent random seeds, and the reported AUC reflects the mean across runs. Sensitivity, specificity, accuracy, and F1-score were evaluated at the Youden’s index-optimal threshold derived from the training cohort and applied unchanged to the external validation cohort. Training performance was estimated using 10-fold stratified cross-validation.

Cause-specific mortality within the CODE-15% dataset was analyzed using a competing risks framework,^16^ in which heart failure-related death was the event of interest and non-heart failure deaths were treated as competing events. Cumulative incidence was estimated using the Aalen-Johansen estimator and between-group differences assessed using Gray’s test. The association between predicted risk group from both models and heart failure-specific mortality was examined using Fine-Gray subdistribution hazard regression, with results reported as subdistribution hazard ratios (SHR) with 95% confidence intervals.

To compare agentic LVDD models with an established ECG-based risk score,^22^ time-dependent discriminative performance for heart failure-related death was assessed using the cumulative/dynamic time-dependent AUC (C/D tdAUC), which evaluates discrimination between patients experiencing the event within a given window and those remaining event-free, accounting for censoring across follow-up. The tdAUC was estimated across time points spanning the 10th to 90th percentile of observed event times and summarized as the mean AUC. Performance of both models was compared against ECG2HF,^22^ with the absolute difference in mean tdAUC (ΔAUC) quantifying incremental gain for each model.

The association between model-predicted phenogroup and guideline-based risk classifications with structural heart disease outcomes was examined using logistic regression, with results reported as odds ratios (OR) with 95% confidence intervals, performed separately for both models across all structural heart disease categories.

All tests were two-sided with statistical significance set at p < 0.05. Statistical analyses were performed in Python (version 3.8.18) and R (version 4.6.0).

## Results

Baseline characteristics of the training and external validation cohorts are presented in **Table 1**. Although the cohorts were broadly similar in age, they differed substantially in their clinical comorbidity and echocardiography profile. High-risk phenogroups were present in 373 patients (38%) compared with 218 (22%) in the derivation cohort, while guideline-defined Grade 2/3 diastolic dysfunction was observed in 174 (20%) versus 103 (11%) patients, respectively. These differences created a stringent test of model generalizability, providing an opportunity to assess performance under clinically relevant dataset shift.

**Table 1.** Baseline Patient Characteristics and Echocardiographic Findings for the Primary Database.

| Variable | Model Training<br>(n=1,011) | External Validation<br>(n=983) | P-value |
| --- | --- | --- | --- |
| **Demographics and Vitals** |  |  |  |
| Age (years) | 58 (44-68) | 58 (35-69) | 0.085 |
| Sex, n (%) | 484 (48) | 517 (53) | 0.043 |
| BMI (kg/m<sup>2</sup>) | 29 (25-34) | 28 (24-32) | <0.0001 |
| HR (beats/min) | 69 (60-78) | 72 (62-82) | <0.0001 |
| **Risk Factors** |  |  |  |
| DM, n (%) | 193 (19) | 434 (44) | <0.0001 |
| HTN, n (%) | 565 (56) | 495 (50) | <0.01 |
| HLD, n (%) | 560 (56) | 281 (29) | <0.0001 |
| Obesity, n (%) | 436 (43) | 318 (32) | <0.0001 |
| Tobacco use, n (%) | 229 (23) | 250 (25) | <0.0001 |
| CKD, n (%) | 21 (3) | 205 (21) | <0.0001 |
| **Echocardiographic Measurements** |  |  |  |
| EF (%) | 64 (58-68) | 57 (53-61) | <0.0001 |
| E (cm/s) | 0.70 (0.60-0.90) | 0.80 (0.70-0.95) | <0.0001 |
| E/A ratio | 1.00 (0.80-1.30) | 1.20 (0.90-1.60) | <0.0001 |
| A (cm/s) | 0.70 (0.60-0.80) | 0.60 (0.50-0.80) | <0.0001 |
| e' (cm/s) | 9.00 (7.10-11.10) | 8.60 (6.50-11.20) | 0.025 |
| E/e' ratio | 7.90 (6.50-10.07) | 9.30 (7.30-12.60) | <0.0001 |
| TRV (m/s) | 2.23 (2.00-2.50) | 2.30 (2.10-2.60) | <0.0001 |
| LAVI (mL/m<sup>2</sup>) | 23 (18-30) | 29 (23-37) | <0.0001 |
| LVMi (g/m<sup>2</sup>) | 76 (62-93) | 78 (65-95) | <0.01 |
| **Echocardiography-based LVDD Frameworks** |  |  |  |
| High-Risk<br>Phenogroup | 218 (22) | 373 (38) | <0.0001 |
| Guideline-based<br>Grades (II/ III) | 103 (11) | 174 (20) | <0.0001 |

BMI, body mass index; HR, heart rate; DM, diabetes mellitus; HTN, hypertension; HLD, hyperlipidemia; CKD, chronic kidney disease; EF, left ventricular ejection fraction; E, early mitral inflow velocity; A, late mitral inflow velocity; e', early diastolic mitral annular velocity; TRV, tricuspid regurgitation velocity; LAVI, left atrial volume index; LVMi, left ventricular mass index. Data are presented as median (IQR) or n (%).

### ECG-Based Agentic Auto-Discovery of Diastolic Dysfunction

To evaluate whether agentic auto-discovery can assess diastolic dysfunction directly from the 12-lead ECG, the ECG-based model was assessed across both classification frameworks (**Figure 2a; Supplemental Table 3**). Under the phenogroup framework, the ECG-based model achieved a cross-validation AUC of 0.82 ± 0.05, with sensitivity of 60.9 ± 22.9%, specificity of 81.4 ± 13.9%, F1-score of 53.6 ± 11.5%, and accuracy of 76.7 ± 7.2%. On the external validation cohort, the model achieved an AUC of 0.87 (95% CI: 0.85-0.89), with sensitivity of 78.6%, specificity of 81.6%, F1-score of 75.3%, and accuracy of 80.5%.

**Figure 1.**
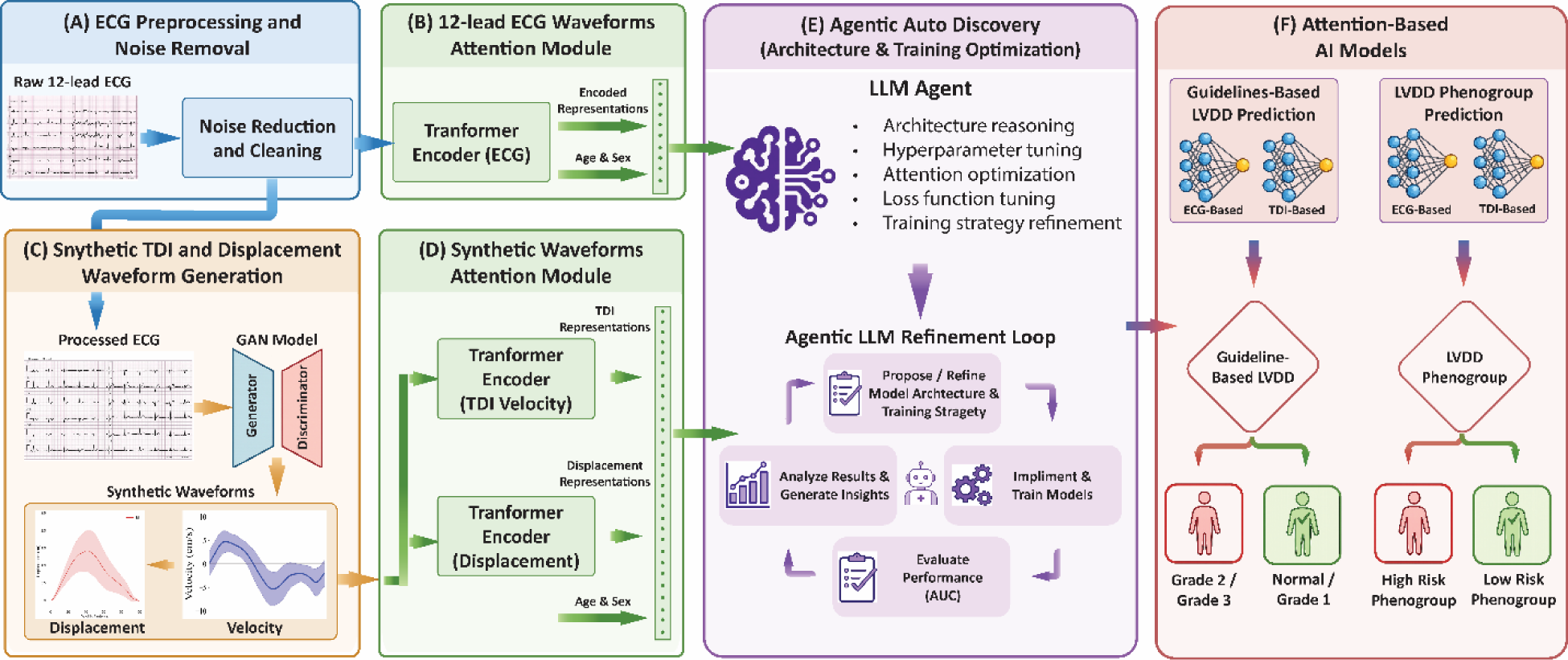
Model Development Pipeline for Agentic Auto-Discovery of Diastolic Phenotypes from the 12-Lead ECG. Raw ECGs underwent preprocessing **(A)**, transformer-based encoding **(B)**, and generative AI synthesis of synthetic TDI waveforms **(C)**, encoded through dedicated attention modules **(D)**. An LLM-driven agentic refinement loop optimized model architecture **(E)**, yielding ECG-based and TDI-based models evaluated under phenogroup and guideline-based LVDD classification frameworks **(F)**. ECG, electrocardiogram; LLM, large language model; TDI, tissue Doppler imaging.

**Figure 2.**
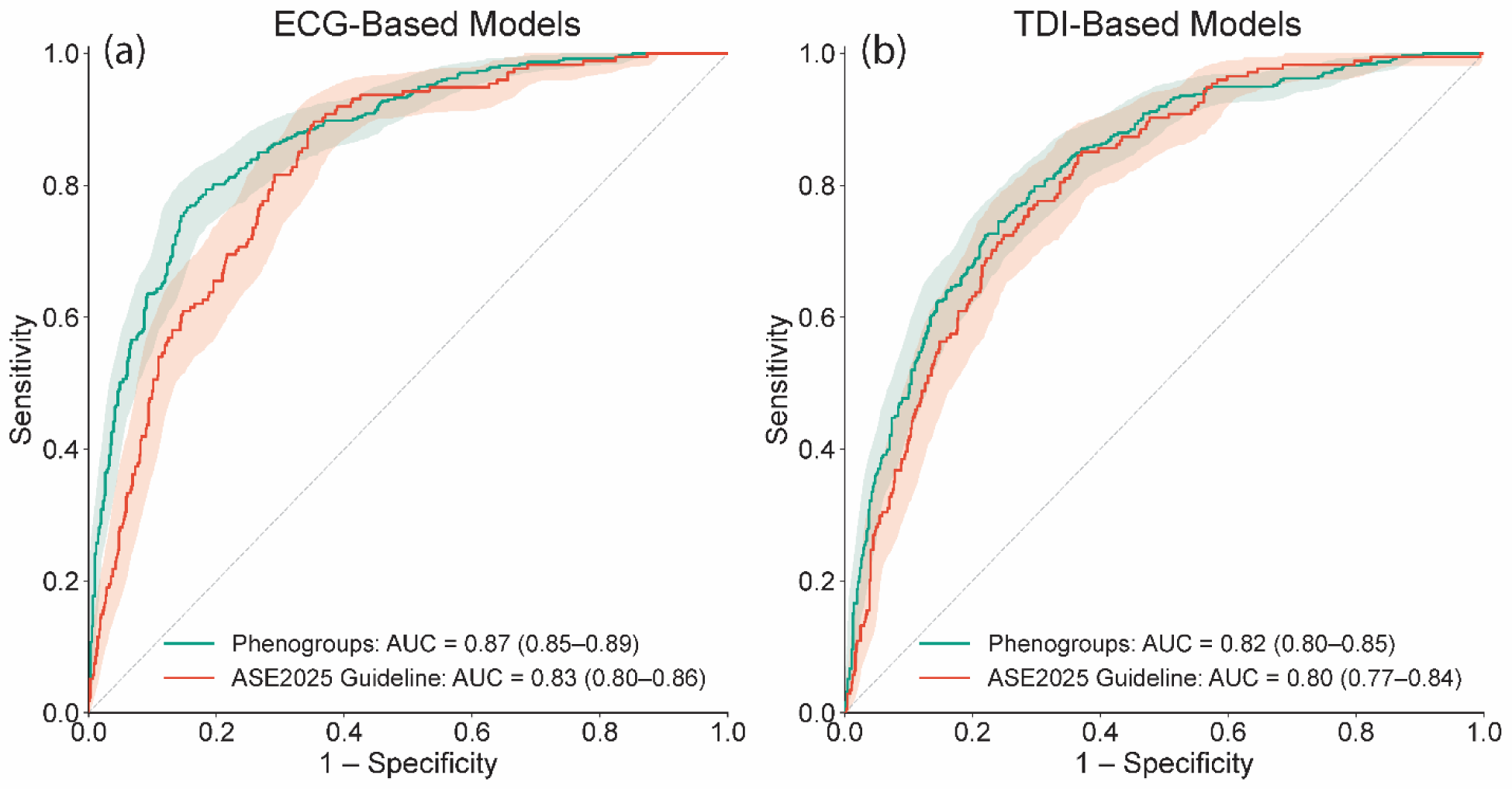
Diastolic Phenotype Classification Performance of ECG-Based and TDI-Based Models. ROC curves are shown for the ECG-based model **(a)** and TDI-based model **(b)** under both the phenogroup (teal) and guideline (red) classification frameworks on the external validation cohort. Shaded areas represent 95% confidence intervals. AUC, area under the receiver operating characteristic curve; CI, confidence interval; TDI, tissue Doppler imaging.

Under the guideline framework, 32 (3.2%) and 97 (9.9%) patients in the training and external validation cohorts respectively were classified as indeterminate and excluded. The ECG-based model achieved a cross-validation AUC of 0.83 ± 0.04, with sensitivity of 69.5 ± 17.2%, specificity of 75.6 ± 20.9%, F1-score of 40.5 ± 8.9%, and accuracy of 75.1 ± 17.1%. On the external validation cohort, the model achieved an AUC of 0.83 (95% CI: 0.80-0.86), with sensitivity of 63.8%, specificity of 81.0%, F1-score of 52.9%, and accuracy of 77.7%.

### TDI-Based Agentic Auto-Discovery of Diastolic Dysfunction

To evaluate whether ECG-derived synthetic TDI waveforms independently support diastolic phenotype classification, the TDI-based model was assessed across both classification frameworks (**Figure 2b; Supplemental Table 3**). Under the phenogroup framework, the TDI-based model achieved a training AUC of 0.79 ± 0.05, with sensitivity of 61.3 ± 13.6%, specificity of 80.4 ± 7.8%, F1-score of 53.8 ± 5.6%, and accuracy of 76.0 ± 4.0%. On the external validation cohort, the model achieved an AUC of 0.82 (95% CI: 0.80-0.85), with sensitivity of 54.7%, specificity of 88.0%, F1-score of 62.8%, and accuracy of 75.4%.

Under the guideline framework, the TDI-based model achieved a training AUC of 0.82 ± 0.05, with sensitivity of 64.8 ± 19.6%, specificity of 77.4 ± 11.7%, F1-score of 36.9 ± 10.0%, and accuracy of 76.1 ± 9.4%. On the external validation cohort, the model achieved an AUC of 0.80 (95% CI: 0.77-0.84), with sensitivity of 24.7%, specificity of 95.8%, F1-score of 34.8%, and accuracy of 81.8%.

### HF-Related Mortality Risk Stratification

Of 219,567 patients, 10,651 (4.85%) died during follow-up: 882 (0.40%) from heart failure, 789 (0.36%) from myocardial infarction, 854 (0.39%) from other cardiac causes, and 8,126 (3.70%) from non-cardiac causes. Both models demonstrated significant separation of HF-related mortality risk across both classification frameworks (all Gray’s p<0.001; **Figure 3**). For the ECG-based model, the phenogroup and guideline-based classifications were independently associated with HF-related mortality (SHR 8.8, 95% CI 7.6-10.2 and SHR 9.5, 95% CI 8.2-11.0, respectively; all p<0.0001). Corresponding SHR values for the TDI-based model were 8.5 (95% CI 7.1-10.1) and 5.5 (95% CI 4.8-6.3), respectively (all p<0.0001; **Supplemental Table 4**). These findings demonstrate robust and clinically meaningful HF-related mortality stratification across both classification frameworks.

**Figure 3.**
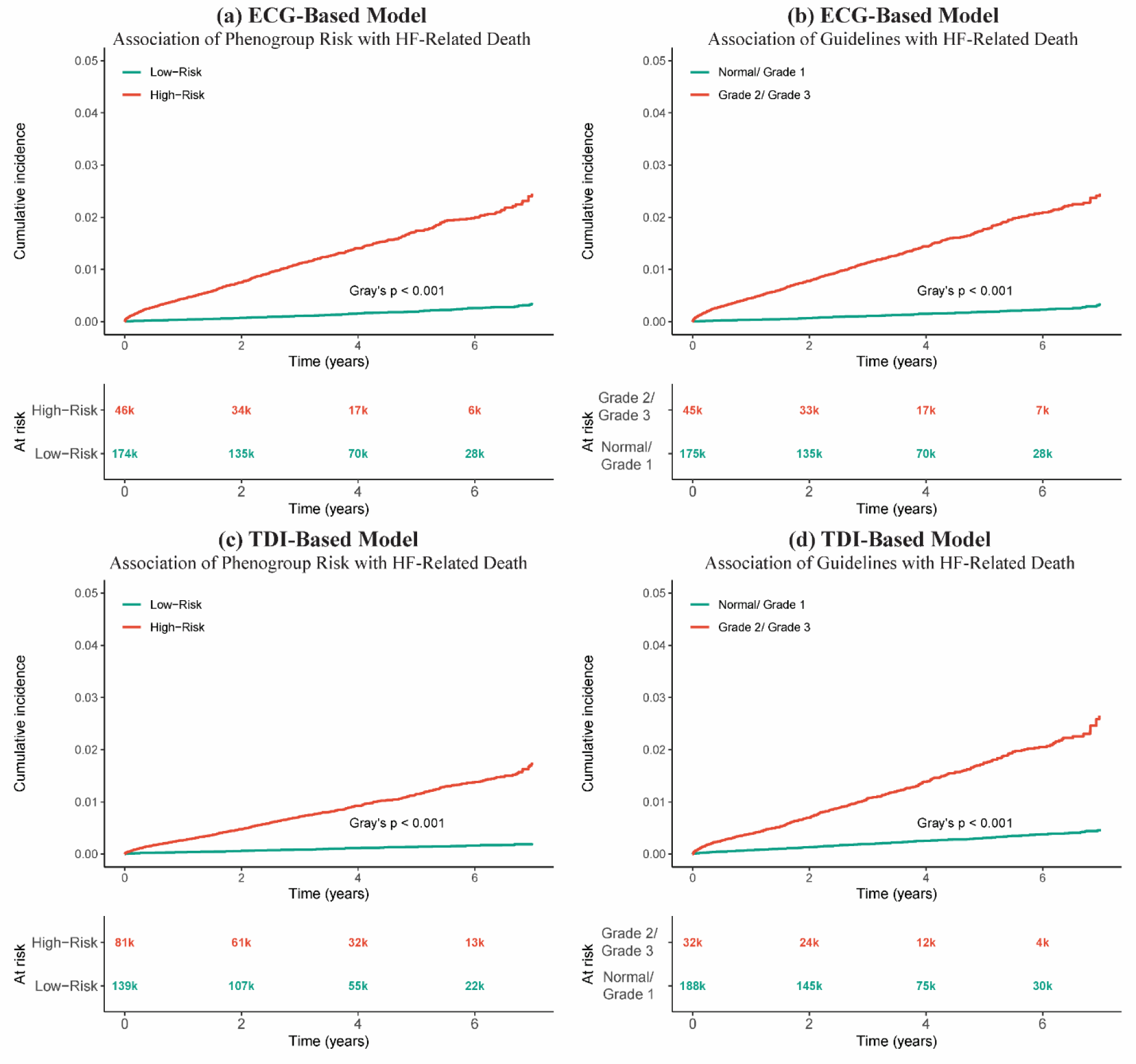
Cumulative Incidence of Heart Failure-Related Mortality Stratified by Model-Predicted Risk Groups in the CODE-15% Cohort (n=219,567). Competing risks-adjusted cumulative incidence functions are shown for the ECG-based model under phenogroup **(a)** and guideline-based **(b)** classification, and for the TDI-based model under phenogroup **(c)** and guideline-based **(d)** classification. All comparisons demonstrated significant separation between risk groups (Gray’s p<0.001). Numbers at risk are shown below each panel. HF, heart failure; TDI, tissue Doppler imaging.

### Time-Dependent Discrimination and Comparison with ECG2HF

To determine whether phenogroup and guideline-based risk classifications could discriminate HF-related death over time, both models were benchmarked against ECG2HF (mean AUC: 0.65 ± 0.02)—a model trained specifically to predict heart failure from the ECG (N=219,443; Events=887).

The ECG-based model achieved a mean time-dependent AUC of 0.86 ± 0.00 under the phenogroup framework (ΔAUC +0.20 over ECG2HF; **Figure 4a**) and 0.86 ± 0.01 under the guideline framework (ΔAUC +0.20 over ECG2HF; **Figure 4b**), demonstrating stable discrimination across the entire five-year follow-up period.

**Figure 4.**
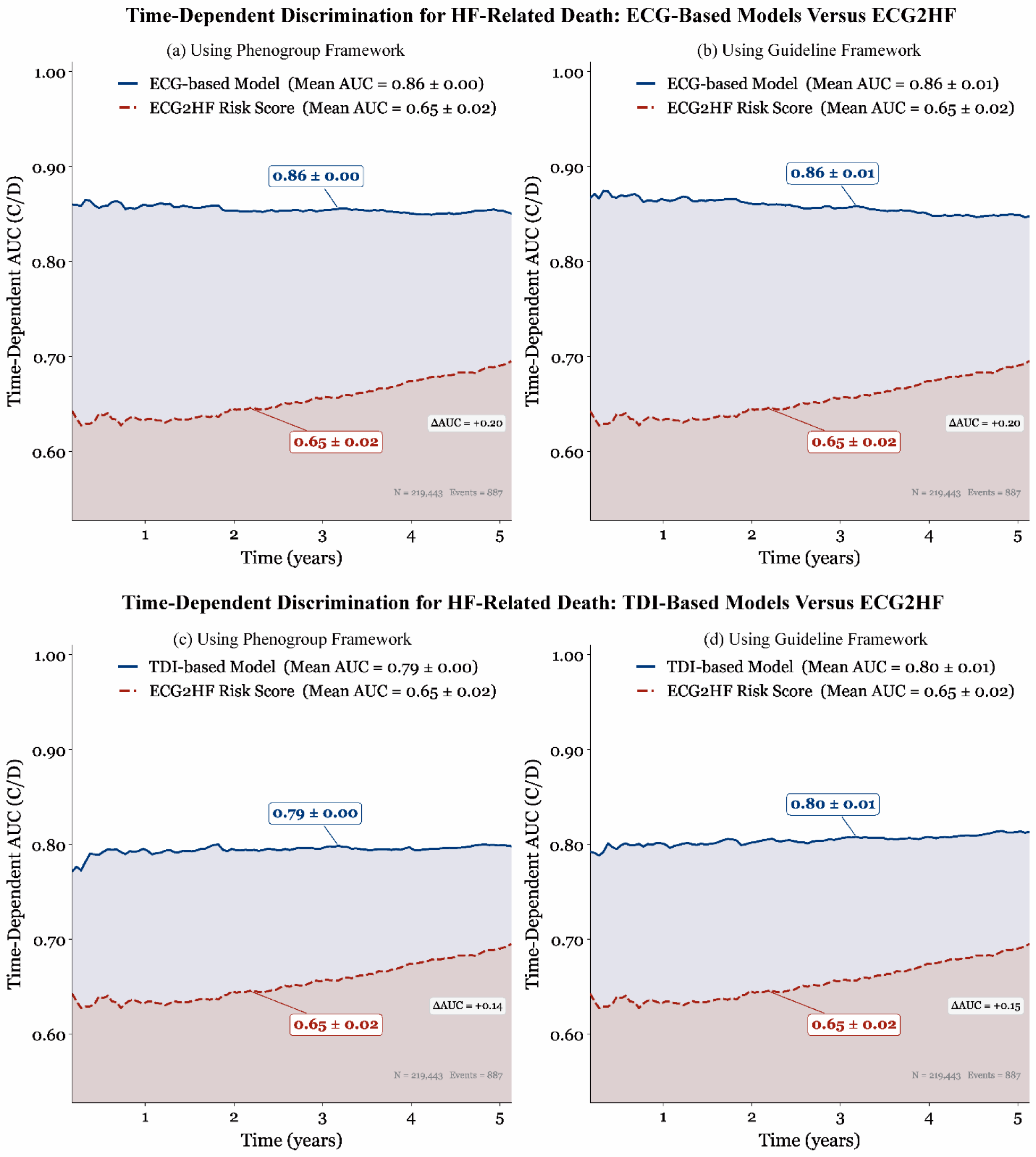
Time-Dependent Discrimination of Diastolic Dysfunction Models for Heart Failure Mortality Against the ECG2HF Risk Score. Time-dependent AUC (cumulative/dynamic estimator) over five years is shown for the ECG-based model using phenogroup **(a)** and guideline **(b)** frameworks, and the TDI-based model using phenogroup **(c)** and guideline **(d)** frameworks, each compared against ECG2HF (red dashed line). Mean AUC values and ΔAUC relative to ECG2HF are displayed within each panel. AUC, area under the receiver operating characteristic curve; C/D, cumulative/dynamic; ECG, electrocardiogram; TDI, tissue Doppler imaging; ΔAUC, difference in mean time-dependent AUC.

The TDI-based model achieved a mean time-dependent AUC of 0.79 ± 0.00 under the phenogroup framework (ΔAUC +0.14 over ECG2HF; **Figure 4c**) and 0.80 ± 0.01 under the guideline framework (ΔAUC +0.15 over ECG2HF; **Figure 4d**), similarly demonstrating stable discrimination across follow-up.

### Associations with Structural and Functional Cardiac Abnormalities

Of 35,718 patients in the EchoNext cohort, the prevalence of echocardiographically confirmed structural heart disease was: LVH 18.8%, LV systolic dysfunction 20.4%, RV systolic dysfunction 7.9%, PASP ≥45 mmHg 12.8%, valvular heart disease 15.2%, and any structural heart disease 43.2%. Both models demonstrated significant associations with all six structural heart disease categories across both classification frameworks (**Figure 5**). Associations were directionally consistent across ECG-based and TDI-based models and across phenogroup and guideline-based classifications. Full OR values for all conditions are presented in **Figure 5**. Valvular heart disease consistently showed the strongest association (OR range 3.5-4.1), followed by pulmonary hypertension (OR range 2.3-2.6) and any structural heart disease (OR range 1.8-2.6).

**Figure 5.**
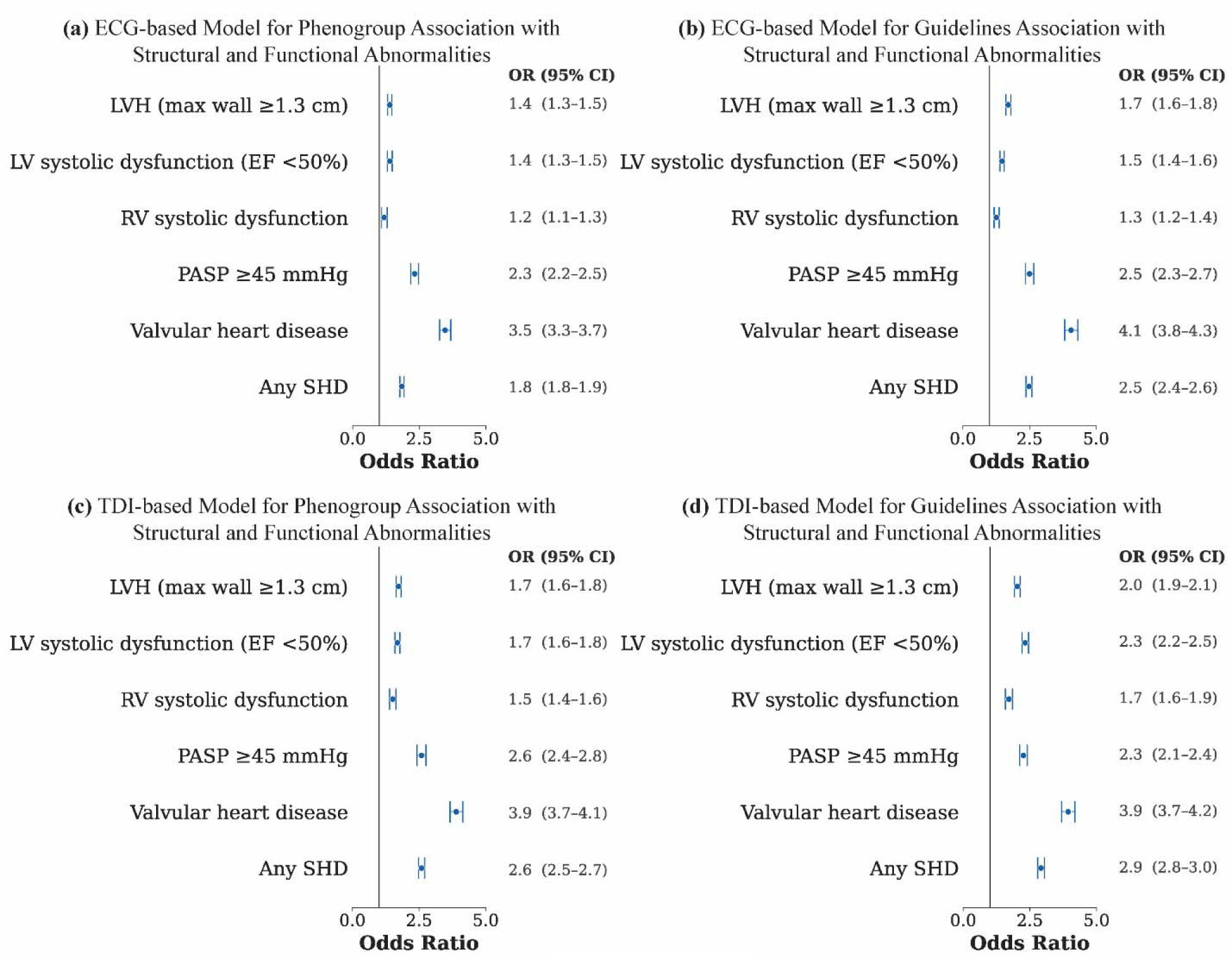
Association of Model-Predicted Risk Classifications with Structural and Functional Cardiac Abnormalities (EchoNext, n=35,718). Odds ratios (95% CI) for the ECG-based model under phenogroup **(a)** and guideline **(b)** frameworks, and the TDI-based model under phenogroup **(c)** and guideline **(d)** frameworks. LVH, left ventricular hypertrophy; LV, left ventricle; RV, right ventricle; PASP, pulmonary artery systolic pressure; SHD, structural heart disease; OR, odds ratio; CI, confidence interval.

## Discussion

Despite its central role in heart failure pathophysiology, diastolic dysfunction assessment remains dependent on multiparametric echocardiography—a resource-intensive modality unavailable at the point of initial clinical contact in most settings. The data-driven phenogroup labels and guideline-based LVDD grades used as reference standards in this study were derived from a comprehensive multiparametric echocardiographic analysis.^4–6^ Together these approaches capture complementary dimensions of diastolic dysfunction, with phenogrouping emphasizing data-driven physiological similarity and guideline-based grading reflecting contemporary expert-defined clinical classification. The central question addressed here is whether these labelled groups, originally discovered in full echo space, can be recovered from the ECG either through direct ECG waveform analysis or through ECG-derived synthetic TDI waveforms that approximate key echocardiographic signals without requiring echo acquisition.^10^ The principal finding is that both pathways successfully identified diastolic phenotypes, suggesting meaningful encoding of echocardiographic information into the ECG signal.

Prior ECG-based approaches to diastolic assessment relied on hand-crafted features or fixed architectures,^8,9,15^ which may under- or over-fit specific datasets and limit generalizability across clinical sites.^23^ The agentic auto-discovery framework addresses recognized limitations of conventional AI development by introducing autonomous architectural optimization tailored specifically to cardiac waveform analysis. The agentic framework autonomously proposes, evaluates, and refines candidate architectures, enabling systematic exploration of a substantially larger design space than is practical through manual development alone. By iteratively learning from prior model performance, the framework can identify non-obvious architectural solutions that may be overlooked by conventional approaches while reducing investigator-dependent design bias. Importantly, this process shifts human effort from model engineering toward defining clinically meaningful objectives and evaluating biological plausibility. In the present study, this enabled robust identification of diastolic dysfunction from both electrical and synthetic mechanical representations of cardiac physiology despite a relatively modest training sample with substantial differences between derivation and validation cohorts. Beyond performance, agentic systems may reduce technical barriers to entry, accelerate hypothesis generation, and facilitate the development of clinically useful models from increasingly complex biomedical datasets.

Both models significantly stratified heart failure-related mortality risk in a large external cohort. Notably, phenogroup-based risk stratification yielded similar subdistribution hazard ratios between the ECG-based and TDI-based models (SHR: 8.8 vs 8.5). This is consistent with prior work demonstrating that machine learning-derived diastolic phenotypes provide independent outcome stratification value compared with guideline-based classification,^24,25^ and supports a broader shift from rule-based staging toward data-driven phenotype-driven risk assessment in diastolic dysfunction. The difference in guideline-based SHRs between models (9.5 vs 5.5) was related to differences in Youden’s index-derived classification thresholds between frameworks rather than a fundamental difference in discriminative capacity.

The comparison of diastolic dysfunction classifications against ECG2HF provides novel context for the incremental value of diastolic phenotyping beyond established ECG-based heart failure risk scoring.^22^ Despite being trained to classify diastolic phenotypes rather than predict heart failure directly, both models exceeded ECG2HF across both classification frameworks, with mean time-dependent AUCs of 0.86 and 0.79-0.80 for the ECG-based and TDI-based models respectively, compared to 0.65 for ECG2HF. Furthermore, both models demonstrated stable discrimination across the entire five-year follow-up, whereas ECG2HF showed a gradual increase in performance over time. This suggests that diastolic phenotype classifications capture a qualitatively different and earlier cardiac risk signal than conventional heart failure risk scoring, consistent with evidence that subclinical diastolic dysfunction precedes clinical heart failure by years.^2,26^

Diastolic dysfunction is not a single disease entity but rather the common physiological consequence of diverse cardiovascular abnormalities, including myocardial hypertrophy, fibrosis, ischemia, valvular heart disease, and chamber remodeling, each of which may contribute distinct structural and functional signatures. Both models demonstrated significant associations with a broad spectrum of structural and functional cardiac abnormalities in an independent echocardiographic validation cohort. Valvular heart disease showed the strongest association across both models and both classification frameworks—an expected finding given the well-established mechanistic relationship between valvular lesions, elevated left ventricular filling pressures, and diastolic impairment.^3,5^ These findings are consistent with prior work demonstrating that AI-enabled ECG analysis can identify a wide range of structural heart disease conditions, ^18^ and extend these observations to ECG-derived diastolic phenotypes.

Collectively, these findings support a broader clinical vision for ECG-based diastolic phenotyping as a scalable, echocardiography-independent approach to cardiac risk assessment. The 12-lead ECG is ubiquitous, inexpensive, and requires no specialist expertise, making it an ideal platform for opportunistic diastolic screening at the point of first clinical contact—in primary care, emergency departments, and pre-operative settings—where echocardiography is unavailable.^27^ Earlier identification of high-risk individuals may facilitate targeted imaging and preventive intervention. Beyond risk stratification, the synthetic TDI pathway offers an additional clinical advantage by providing a physiologically interpretable waveform alongside risk classification.^10^

### Limitations and opportunities

Several limitations warrant consideration. First, both models were developed in tertiary care populations, which may limit generalizability to community settings with lower disease prevalence, although the large-scale external validation in the CODE-15% primary care cohort partially mitigates this concern. Second, both guideline-based grading and phenogroup assignment were used as reference standards rather than invasive measurements of left ventricular filling pressures. Although both frameworks have been independently validated and are associated with clinically meaningful outcomes, some degree of misclassification is inherent to any noninvasive assessment of diastolic function. In addition, disease-specific guideline algorithms were not applied in certain populations, such as patients with atrial fibrillation or specific structural heart diseases within the EchoNext cohort. Instead, the same ECG-based agentic models were applied uniformly across the entire dataset, which may have introduced additional classification error in these subgroups. Third, patients with arrhythmias at the time of ECG acquisition were excluded from model development; therefore, performance in populations with atrial fibrillation and other rhythm disturbances remains incompletely characterized. Nevertheless, both the CODE-15% and EchoNext validation cohorts included a modest number of patients with atrial fibrillation, providing preliminary evidence that the models retain utility in the presence of rhythm disturbances. Fourth, the analyses were based on single time-point ECG and echocardiographic assessments. Whether model outputs track longitudinal changes in diastolic function, treatment response, or progression to heart failure will require prospective investigation. Fifth, diastolic dysfunction represents a heterogeneous syndrome arising from diverse underlying pathophysiological processes. Although the models demonstrated robust performance across clinically distinct cohorts, further studies are needed to evaluate performance within specific disease subgroups and across the spectrum of disease etiologies. Finally, while both models demonstrated meaningful heart failure-related mortality risk stratification, prospective studies are required to determine whether model-guided screening, diagnosis, or therapeutic decision-making can improve patient outcomes.

## Conclusion

Diastolic dysfunction can be classified from the surface 12-lead ECG using attention-based agentic AI, whether applied directly to raw ECG waveforms or to generative AI-derived synthetic TDI waveforms. Both approaches demonstrated meaningful discrimination across data-driven phenogroup and guideline-based classification frameworks, with the TDI-based model independently demonstrating sufficient signal for phenotype classification without echocardiographic acquisition. Both models significantly stratified HF-related mortality risk and demonstrated consistent associations with structural and functional cardiac abnormalities across both classification frameworks. These findings support the ECG as a scalable, echocardiography-independent substrate for diastolic function assessment. Prospective studies evaluating the clinical impact of ECG-based diastolic stratification on patient outcomes and population-scale screening are warranted.

## Funding Source and Acknowledgements

This study was supported by the National Institutes of Health/National Heart, Lung, and Blood Institute (NIH/NHLBI; grant R01HL173998). AJ would like to thank Srinidhi Sunkara, MS, for providing volunteer research support in the development of the agentic model framework and for offering feedback on model assessment.

## Data Availability

The clinical and feature datasets generated and analyzed during the current study are not publicly available due to institutional regulations and the terms of our ethics/IRB approval, which do not permit open sharing of individual-level patient data. De-identified data underlying the main results may be made available from the corresponding author upon reasonable request and subject to institutional approvals and a data use agreement.

---

**Central Illustration. Agentic Auto-Discovery of Diastolic Phenotypes from the 12-Lead ECG.** Synthetic TDI waveforms were generated from paired ECGs using a generative AI model. ECG-based and TDI-based agentic AI models were developed and assessed for HF-related mortality risk stratification and structural heart disease associations, benchmarked against ECG2HF.

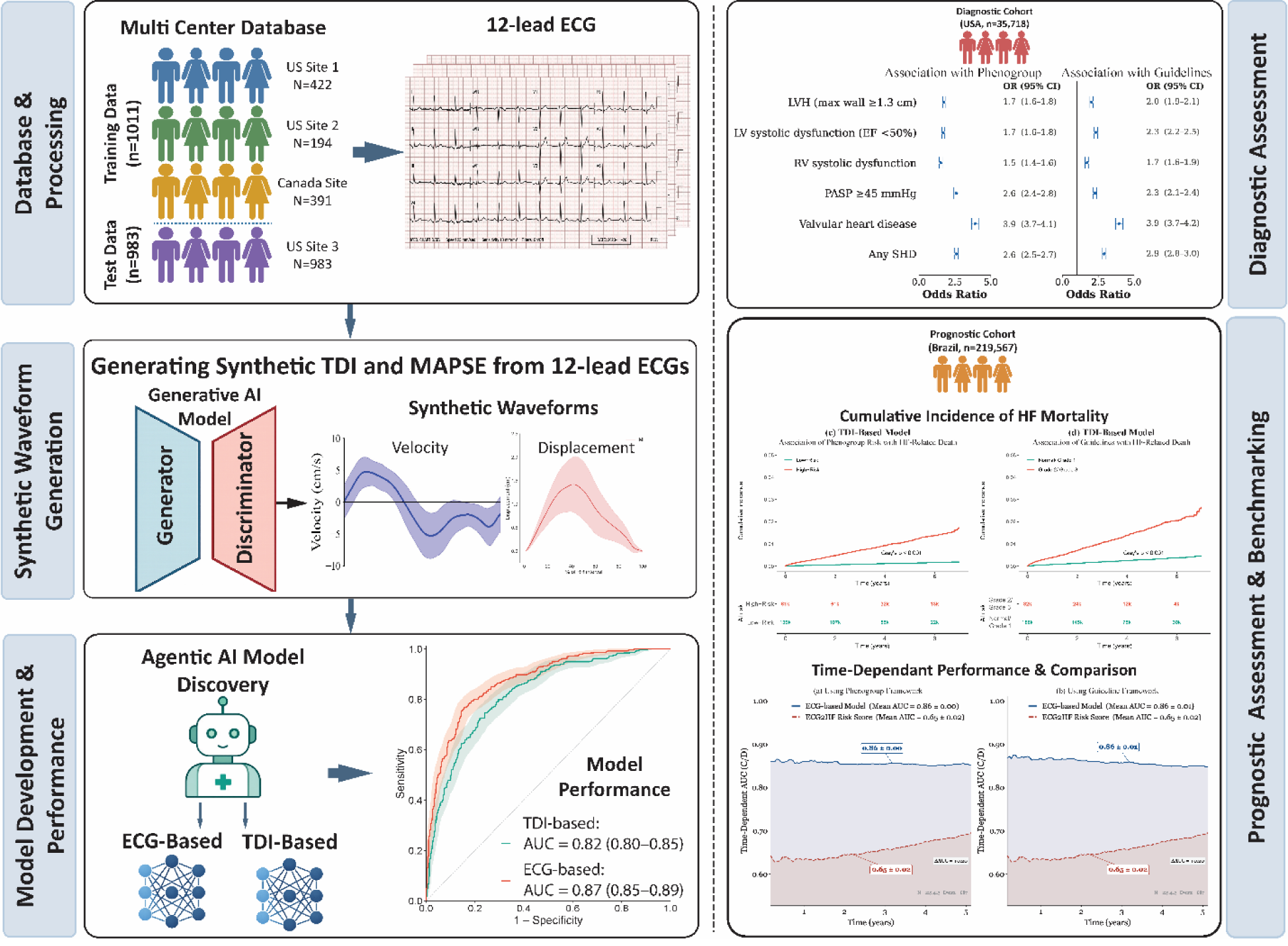

