## Supplemental Material for "Agentic Autodiscovery of Diastolic Dysfunction Phenotypes from Surface Electrocardiogram"

### S1. ECG Acquisition and Preprocessing—Technical Details

Twelve-lead ECGs were recorded and interpreted using the University of Glasgow Interpretive Analysis program (release 28.5, January 2014). All ECGs were exported from MUSE NX (GE Healthcare) as XML files, de-identified, and stored as CSV files. Raw signals were converted to millivolts using appropriate scaling factors and underwent band-pass filtering (0.5–40 Hz) and quality control. Waveforms were segmented from R-peak to R-peak, normalized to a canonical 512-sample window via linear interpolation, and smoothed using a moving average filter (window size = 3). For the EchoNext dataset, only the first recorded ECG per unique patient was retained. For the CODE-15% dataset, individual patient files containing raw 12-lead ECG tracings sampled at 400 Hz were used alongside an accompanying exam.csv file providing clinical data and follow-up survival information.

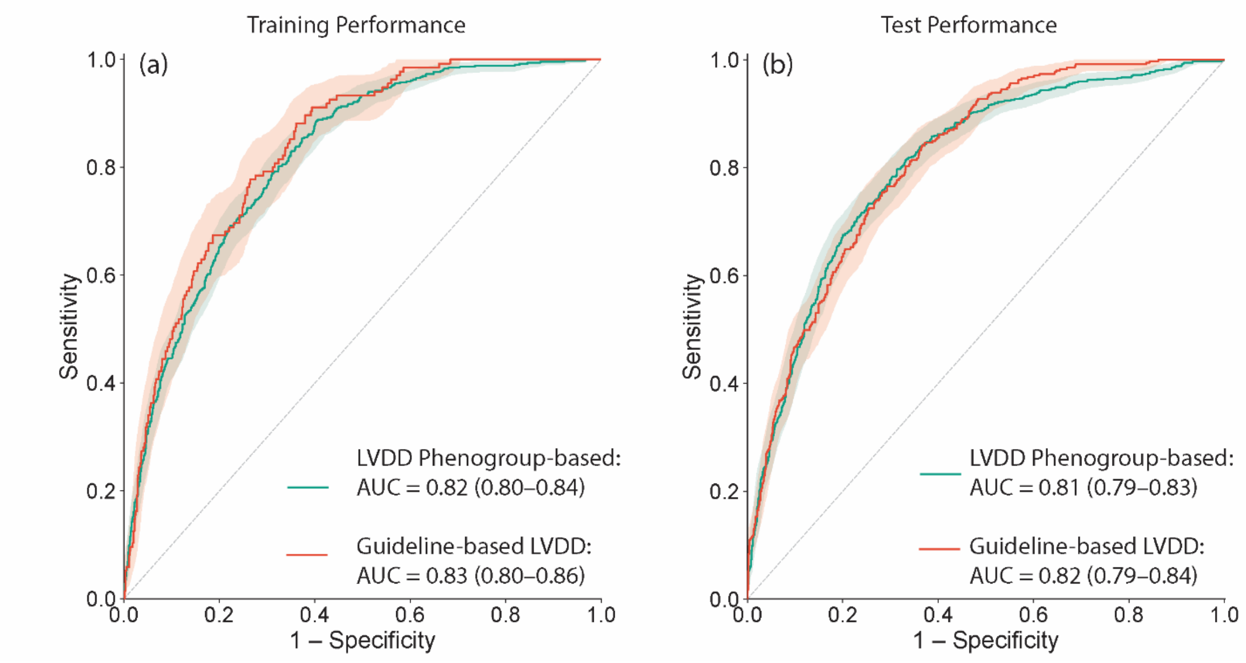


**Supplemental Figure 1.** Receiver Operating Characteristics Performance of a Convolutional Neural Network Using Real Echocardiographic TDI Waveforms for LVDD Classification and Phenogroup Prediction: Establishing the Mechanistic Rationale for the Synthetic TDI Approach.

**Supplemental Table 1.** Diagnostic Performance of a Convolutional Neural Network Using Real Echocardiographic TDI Waveforms for LVDD Classification and Phenogroup Prediction: Establishing the Mechanistic Rationale for the Synthetic TDI Approach.

| | LVDD Phenogroup | | Guideline-Based LVDD | |
|---|---|---|---|---|
| **Metric** | **Training** | **Test** | **Training** | **Test** |
| Sensitivity (%) | 74.2±5.7 | 73.5 | 68.1±9.4 | 66.4 |
| Specificity (%) | 71.9±4.6 | 79.5 | 78.7±4.7 | 77.2 |
| F1 Score (%) | 52.9±3.2 | 73.5 | 35.2±5.7 | 51.1 |
| Accuracy (%) | 72.4±3.2 | 75.0 | 77.8±4.1 | 75.1 |
| AUROC | 0.8±0 | 0.81 (0.79–0.83) | 0.8±0 | 0.82 (0.79–0.84) |

A convolutional neural network with an InceptionNet architecture was trained on directly acquired TDI waveform images to provide a reference benchmark for comparison with the ECG-derived synthetic TDI-based and ECG-based agentic AI models. Model development used 10-fold cross-validation; training metrics reflect mean ± standard deviation across folds. Test metrics reflect performance on the held-out external validation cohort with 95% confidence intervals reported for AUROC. This table serves as a reference standard representing the performance ceiling achievable using real echocardiographically acquired TDI waveforms.

**S2. Model Architecture Specifications**

*Overview*

Both model configurations shared the same underlying architecture, in which each input signal is processed through a dedicated encoder, representations are combined through cross-modal attention layers, and a final classification layer produces a probability of high-risk diastolic dysfunction. Three encoders were used:

(i) Waveform encoder: processes each synthetic TDI waveform sequentially, learning which portions of the cardiac cycle carry the most diagnostic information

(ii) ECG encoder: processes the 12-lead ECG in two stages — within-lead morphology followed by cross-lead relationships

(iii) Demographic encoder: converts age and sex into numerical representations compatible with the signal encoders

When a signal is unavailable, a dedicated learned placeholder embedding — one per input modality, optimized jointly with the rest of the model during training — is substituted in its place, allowing the model to generate predictions from any non-empty combination of available inputs. Layer normalization was applied at all normalization sites throughout the network. Batch normalization was evaluated during the agentic architecture search but excluded because it introduced systematic performance differences between the training and external validation cohorts, arising from differences in data distribution between batches across sites.

*Combinatorial Modality-Masking and Model Training*

The model was trained simultaneously on all possible combinations of input signals—a procedure known as combinatorial modality-masking—ensuring reliable predictions regardless of which inputs are available in clinical use. For a model with k inputs, this generates $2^k-1$ non-empty combinations (ECG-based model: k=3, 7 combinations; TDI-based model: k=4, 15 combinations). When a signal is unavailable, a learned placeholder optimized during training is substituted.

The classification loss was designed to address class imbalance by up-weighting the positive class and down-weighting easily classified negatives, with an additional ranking term aligning training with the primary AUC metric. ECG inputs received three augmentation strategies during training—amplitude variation, additive noise, and circular time-axis shifting—with five augmented forward passes averaged at inference. Each final model was retrained 20 times under independent random initializations; reported AUC reflects the mean across runs with standard deviation quantifying optimization stability.

*Agentic Auto-Discovery—LLM-Driven Refinement Loop*

An automated LLM-driven refinement loop iteratively proposed and tested architectural modifications guided by observed performance, replacing manual tuning with a closed-loop discovery process. Each iteration proceeded through four stages: propose, validate, implement, and accept or revert.

**Supplemental Table 2.** Feasibility of Key Echocardiographic Measurements in the Training and External Validation Cohorts.

| Variable | Feasibility in Training Data (%) | Feasibility in Test Data (%) |
|---|---|---|
| Septal e' (cm/s) | 100 | 99.2 |
| Lateral e' (cm/s) | 99.7 | 75.9 |
| Septal E/e' | 99.3 | 96.5 |
| Lateral E/e' | 99.1 | 74.7 |
| TR Velocity (m/s) | 79.7 | 67.2 |
| E/A | 98.5 | 93.1 |
| LA Strain (%) | 85.3 | 89.4 |
| LAVI (mL/m²) | 98.6 | 87.6 |

Feasibility reflects the proportion of patients with available measurements for each echocardiographic parameter. e′, early diastolic mitral annular velocity; E/e′, ratio of mitral inflow velocity to mitral annular velocity; TR, tricuspid regurgitation; E/A, ratio of early to late mitral inflow velocity; LA, left atrium; LAVI, left atrial volume index.

**Supplemental Table 3**. Diagnostic Performance of the ECG-Based and TDI-Based Models for Guideline-Based LVDD Classification and Data-Driven Phenogroup Prediction Across Training and Test Sets

| | ECG-based Model | | TDI-based Model | |
|---|---|---|---|---|
| **Metric** | **Training** | **Test** | **Training** | **Test** |
| **Phenogroup Prediction** | | | | |
| Threshold | 0.29 | 0.29 | 0.292 | 0.292 |
| AUROC | 0.82 ± 0.05 | 0.87 | 0.79 ± 0.05 | 0.82 |
| Sensitivity | 60.95 ± 22.86 | 78.6 | 61.3 ± 13.6 | 54.7 |
| Specificity | 81.4 ± 13.9 | 81.6 | 80.4 ± 7.8 | 88 |
| F1 Score | 53.6 ± 11.5 | 75.3 | 53.8 ± 5.6 | 62.8 |
| Accuracy | 76.7 ± 7.2 | 80.5 | 76.0 ± 4.0 | 75.4 |
| **Guideline-Based Grade Predictions** | | | | |
| Threshold | 0.109 | 0.109 | 0.136 | 0.136 |
| AUROC | 0.83 ± 0.04 | 0.83 | 0.82 ± 0.05 | 0.80 |
| Sensitivity | 69.5 ± 17.2 | 63.8 | 64.8 ± 19.6 | 24.7 |
| Specificity | 75.6 ± 20.9 | 81.0 | 77.4 ± 11.7 | 95.8 |
| F1 Score | 40.5 ± 8.9 | 52.9 | 36.9 ± 10.0 | 34.8 |
| Accuracy | 75.1 ± 17.1 | 77.7 | 76.1 ± 9.4 | 81.8 |

Performance metrics are reported separately for the training and test sets across both classification frameworks. Training metrics reflect mean ± standard deviation across 20 independent retraining runs under different random seeds, quantifying optimization stability. Test metrics reflect performance on the held-out external validation cohort. The classification threshold for each model was determined using Youden's index applied to the training set and held fixed for test set evaluation. AUROC, area under the receiver operating characteristic curve; LVDD, left ventricular diastolic dysfunction; TDI, tissue Doppler imaging.

**Supplemental Table 4** Prognostic Association of Model-Predicted Risk Groups with Heart Failure–Related Death in the CODE-15% Dataset: Fine-Gray Competing Risks Analysis.

| Classification Framework | Coefficient | SHR (95% CI) | SE | Z | P value |
|---|---|---|---|---|---|
| ECG-Based Model | | | | | |
| Phenogroups | 2.2 | 8.8 (7.6-10.2) | 0.07 | 29.7 | <0.0001 |
| Guidelines | 2.3 | 9.5 (8.2-11.0) | 0.07 | 30.2 | <0.0001 |
| TDI-Based Model | | | | | |
| Phenogroups | 2.1 | 8.5 (7.1-10.1) | 0.09 | 23.5 | <0.0001 |
| Guidelines | 1.7 | 5.5 (4.8-6.3) | 0.07 | 25.4 | <0.0001 |

Fine-Gray subdistribution hazard regression was used to quantify the independent association between model-predicted risk group and heart failure–related death in the presence of competing risks (myocardial infarction–related death, other cardiac death, and non-cardiac death). For guideline-based LVDD prediction, risk groups were defined as Normal/Grade 1 versus Grade 2/Grade 3; for phenogroup-based prediction, groups were defined as Low-Risk versus High-Risk. In both frameworks, thresholds were determined using Youden's index. The predicted risk group was entered as the sole covariate. Subdistribution hazard ratios (SHR) reflect the relative risk of heart failure–related death in the high-risk group compared to the low-risk group, accounting for the subdistribution of competing events. Confidence intervals were computed as $\exp(\beta \pm 1.96 \times SE)$ and p-values derived from the Wald z-statistic.
